# Timing of antiviral treatment initiation is critical to reduce SARS-Cov-2 viral load

**DOI:** 10.1101/2020.04.04.20047886

**Authors:** Antonio Gonçalves, Julie Bertrand, Ruian Ke, Emmanuelle Comets, Xavier de Lamballerie, Denis Malvy, Andrés Pizzorno, Olivier Terrier, Manuel Rosa Calatrava, France Mentré, Patrick Smith, Alan S Perelson, Jérémie Guedj

## Abstract

We modeled the viral dynamics of 13 untreated patients infected with SARS-CoV-2 to infer viral growth parameters and predict the effects of antiviral treatments. In order to reduce peak viral load by more than 2 logs, drug efficacy needs to be greater than 90% if treatment is administered after symptom onset; an efficacy of 60% could be sufficient if treatment is initiated before symptom onset. Given their pharmacokinetic/pharmacodynamic properties, current investigated drugs may be in a range of 6-87% efficacy. They may help control virus if administered very early, but may not have a major effect in severe patients.

## Main text

### Background

The outbreak of severe acute respiratory syndrome coronavirus 2 (SARS-CoV-2), which originated in Wuhan, China, has become a global pandemic. By May 7^th^, 2020, this virus had infected more than 3,700,000 people worldwide and caused more than 260,000 deaths. To readily propose a first line of defense and combat the virus in hospitalized patients, the World Health Organization relies on already existing drugs (“repurposed”) that are immediately available in large quantities and have a good safety profile. In coordination with other European institutions, France is implementing a randomized clinical trial in hospitalized patients (“DisCoVery”, NCT04315948) comparing the efficacy of lopinavir/ritonavir ± IFN-β-1a, remdesivir and hydroxychloroquine in hospitalized patients. However, the clinical efficacy of currently available therapies is unknown and could be limited ^1^.

Here we fit mathematical models of viral dynamics to *in vivo* data to estimate parameters driving viral replication. We then use these models to predict the needed efficacy of treatments ^2^. By combining the expected drug concentrations and EC_50_ of drug candidates, we also use the model to predict the effects of various dosing regimens (doses, timing of treatment initiation) on viral load dynamics.

### Methods

#### Data used for fitting

We used published data from 13 untreated patients infected with SARS-CoV-2 that were followed in 4 Singapore hospitals ^3^. Patients were hospitalized in median at day 3 after onset of symptoms (range: 1-10) and had a median symptomatic period of 12 days (range: 5-24). Viral loads in nasopharyngeal swabs were measured by real time reverse transcriptase polymerase chain reaction (RT PCR, lower limit of quantification: 38 cycles, CT) at multiple time points with an observed peak of viral load at day 5 post onset of symptoms (range: 2-27 days). Data presented in CT were transformed to log_10_ copies/mL using a published relationship in Zou et al. ^4^ and the model was fit to the log_10_ viral load. Of note, the transformation from CT to log_10_ copies/mL does not affect the estimates of parameters of interest, in particular R_0_ and the death rate of productively infected cells. Time since infection was assumed to be 5 days before the onset of symptoms ^5^. In a sensitivity analysis, we also examined values of 2 and 10 days.

#### Model

Viral dynamics was fitted using a target cell limited model with an eclipse phase.

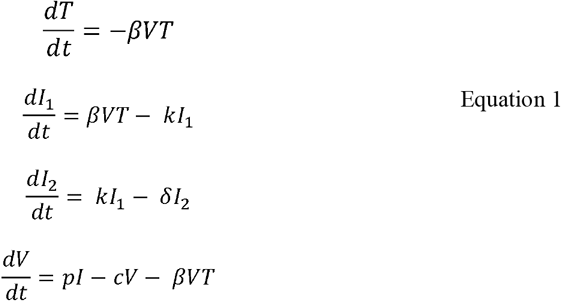

The model considers three populations of cells: target cells, *T*, infected cells in the eclipse phase, *I*_*1*_, and productively infected cells, *I*_*2*._ Given the timescale of the infection, we neglect target cell proliferation and natural death, and we focused on the process of cell depletion by virus infection. We assumed target cells become infected with rate constant *β*. After an average time of 1/*k*, these cells start producing virus and are cleared with per capita rate δ. Virions are released from productively infected cells *I*_*2*_ at rate *p* per cell and are cleared from the circulation at per capita rate *c* or lost by infecting a target cell. Based on this model, the basic reproduction number, *R*_0_, the average number of cells infected by a single infected cell at the beginning of the infection, is ^6,7^

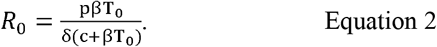

To determine the target cell concentration, the following calculation was done. We assumed that the total number of epithelial cells in the upper respiratory tract was 4 × 10^8^ cells, distributed in a volume of 30 mL ^8^. Assuming that 1% of these cells express the ACE2 receptor and associated proteases needed for viral entry ^9^, the target cell concentration, T_0_, was fixed to 1.33 × 10^5^ cells/mL. Following what was found in other viral infections, including acute infection ^6^, the clearance rate of virus, c, was assumed to be fast and equal to 10 d^-1^ but values of 5 and 20 d^-1^ were also evaluated.

#### Model building strategy

Because not all parameters can be identified when only viral load data are available, parameters V_0_ and k were fixed at 10^-1^ copies/mL and 3 d^-1^, respectively, which corresponds to an initiation of the viral production 8 hours after cell infection on average ^10^. A sensitivity analysis was also performed with different values of k={1, 3, 5}d^-1^ and V_0_={10^×3^, 10^-2^, 10^×1^} copies/mL to assess the robustness of the parameter estimates. The parameter R_0_ was estimated instead of the infection rate *β* by a change of variables in Eq. (1).

Parameters were estimated in a non-linear mixed-effect modeling framework using the SAEM algorithm implemented in Monolix (www.lixoft.com). The model providing the best description of the data was used for the predictions and the individual data fitting, and model averaging was used to correct for the model uncertainty when calculating confidence intervals of estimated parameters ^11^.

#### Predicting the effects of treatment according to the antiviral efficacy and the time of treatment initiation

We assumed that antivirals with a constant effectiveness ε could reduce R_0_ by a factor (1-ε), with ε taking values from 50% to 99% in Equation 2. We considered different times of treatment initiation, from the time of infection to 3 days after the symptom onset. For each treatment strategy, we calculated the reduction in viral load at the peak of infection in the absence of treatment, i.e., 5 days after symptom onset.

#### Model including an innate immune response

We also examined the possibility that cell infection is limited by an innate immune response that renders cells refractory to infection, as was proposed for other acute viral infections ^8,12^. In this model, two additional compartments are added, one for a cytokine (eg., IFN) released in response to antigen, and one representing cells in an antiviral state that cannot be infected (Supplemental information).

#### PK/PD drug properties of lopinavir/ritonavir, hydroxychloroquine, IFN-β-1a and remdesivir

We relied on the literature to find PK population models and parameter values of lopinavir/ritonavir ^13^, plasma hydroxychloroquine ^14^, IFN-β-1a ^15^ and remdesivir as well as reported EC_50_ values *in vitro* (see Table 1). For lopinavir EC_50_, specific results were obtained as follows. Vero E6 cells were infected by SARS-CoV-2 (strain BetaCoV/France/IDF0571/2020) at a MOI of 0.01 and treated with several concentrations of lopinavir one hour after infection. Supernatant samples were collected at 48 and 72 hours post infection. Relative quantification of viral genome was performed by RT-qPCR from RNA extracted using QIAamp viral RNA Mini Kit (Qiagen). IC50 values of lopinavir (5.246 μM and 4.941 μM at 48 and 72 hours post infection, respectively) were calculated from dose-response curve using a four-parameter logistic regression model.

**Table 1:**
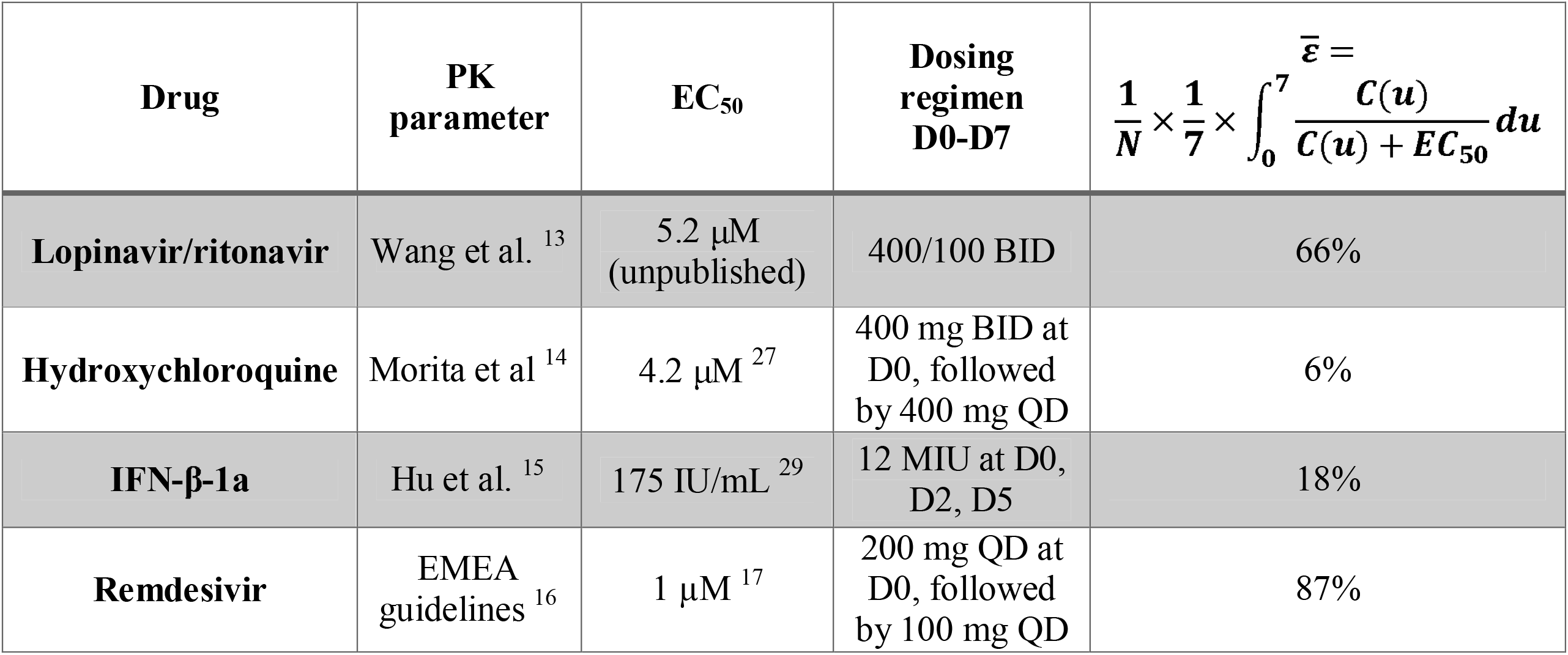
PK/PD properties of candidate antiviral drugs. We assume that the total concentrations were the driver of efficacy, and we did not consider intracellular metabolites or free drug concentrations

To determine the mean antiviral efficacy of these drugs, we simulated their plasma pharmacokinetic profiles considering clinical regimens used in the Discovery trial, namely 400/100 mg twice daily (BID) for lopinavir/ritonavir, 400 mg BID the first day (loading dose) followed by 400 mg once daily (QD) for hydroxychloroquine and 12 MIU for IFN-*β*-1a. For each regimen, we simulated N=100 PK profiles according to the reported parameter distributions. Then, we calculated for each simulated individual the mean inhibitory coefficient, sometimes called the mean antiviral effectiveness,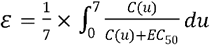 during the first week of treatment, and the mean value over the N profiles was given in Table 1. For comparison purposes, we based the analysis on total plasma concentrations and did not adjust for plasma protein binding when computing efficacy.

Since the pharmacokinetic parameters of remdesivir have not been reported yet in humans, the same method could not be applied. We used the summary statistics reported in the summary for compassionate use of remdesivir filed by Gilead to the EMEA ^16^ to derive the mean concentration of the active metabolite in PBMC, C_mean_, using the AUC after a loading dose of 200 mg and after repeated doses of 100 mg QD and we assumed that the EC_50_ for the metabolite was equal to the EC_50_ of the parent ^17^.

## Results

We used a “target-cell limited” model with an eclipse phase ^8^ given by Eq. (1) to characterize the viral load dynamics of 13 hospitalized patients in Singapore for which data obtained from frequent nasopharyngeal swabs were available ^3^. Because this model needs to incorporate a date of infection, an incubation period of 5 days was used as the most plausible date of infection in each patient ^5^. The model fitted the data well (Fig. 1) and using a model averaging approach to take into account model uncertainty ^11^, the within-host basic reproductive number, R_0,_ was found equal to 8.6 (CI_95%_ = [1.9-17.6]), and the death rate of productively infected cells was estimated as 0.60 d^-1^ (CI_95%_ = [0.22-0.97]), corresponding to a median half-life of 1.2 days (Table 2 and Fig. S1). In influenza A, another respiratory infectious disease, estimates of the within host R_0_ varied greatly, but the half-life of infected cells was shorter than 10 hours (see more details in ^18^), suggesting a faster clearance of influenza infected cells than SARS-CoV-2. The viral production rate p was also estimated as 22.7 copies.d^-1^ (CI_95%_ = [0,59.6]) (Table 2). However, as shown previously, p cannot be uniquely identified unless the initial target cell density T_0_ is known ^19,20^. Therefore, the only quantity that can be reliably estimated is the product p×T_0,_ equal to 3.0 × 10^6^ copies d^-1^ (CI_95%_ = [0-7.9×10^6^]). Parameter estimates and confidence intervals were also consistent across models assuming a viral clearance c of 5 or 20 d^-1^ (see Table S1).

**Table 2:** Median and confidence intervals of R_0_, *δ* and p across models and following model averaging procedure

| **k (d<sup>-1</sup>)** | **V<sub>0</sub>**<br>**(log<sub>10</sub> cp/mL)** | **R<sub>0</sub> [CI<sub>95%</sub>]** | **δ [CI<sub>95%</sub>] (d<sup>-1</sup>)** | **p [CI<sub>95%</sub>] (d<sup>-1</sup>)** |
| --- | --- | --- | --- | --- |
| **1** | **10<sup>-1</sup>** | 13.1 [5.1-21.8] | 0.68 [0.44-0.9] | 26.55 [0-57.32] |
|  | **10<sup>-2</sup>** | 15.4 [7.9-23.2] | 0.71 [0.46-0.94] | 32 [8.38-56.26] |
|  | **10<sup>-3</sup>** | 19.1 [10.3-28.8] | 0.71 [0.46-0.94] | 35.18 [12.46-60.06] |
| **3** | **10<sup>-1</sup>** | 8.2 [3.5-13.1] | 0.6 [0.38-0.82] | 21.36 [0-60.35] |
|  | **10<sup>-2</sup>** | 9.8 [4.1-15.4] | 0.58 [0.08-1.15] | 20.37 [0-85.01] |
|  | **10<sup>-3</sup>** | 12.5 [4.8-20] | 0.58 [0.42-0.74] | 23.37 [6.36-39.57] |
| **5** | **10<sup>-1</sup>** | 7.1 [0-13.9] | 0.6 [0.19-1.06] | 22.07 [0-58.57] |
|  | **10<sup>-2</sup>** | 8.9 [4.7-13.2] | 0.57 [0.43-0.74] | 22.42 [1.9-41.84] |
|  | **10<sup>-3</sup>** | 10.2 [4.8-16.2] | 0.58 [0.46-0.71] | 22.31 [5.48-41.46] |
| **Model averaging** |  | 8.6 [1.9-17.6] | 0.6 [0.22-0.97] | 22.71 [0-59.64] |

**Figure 1:**
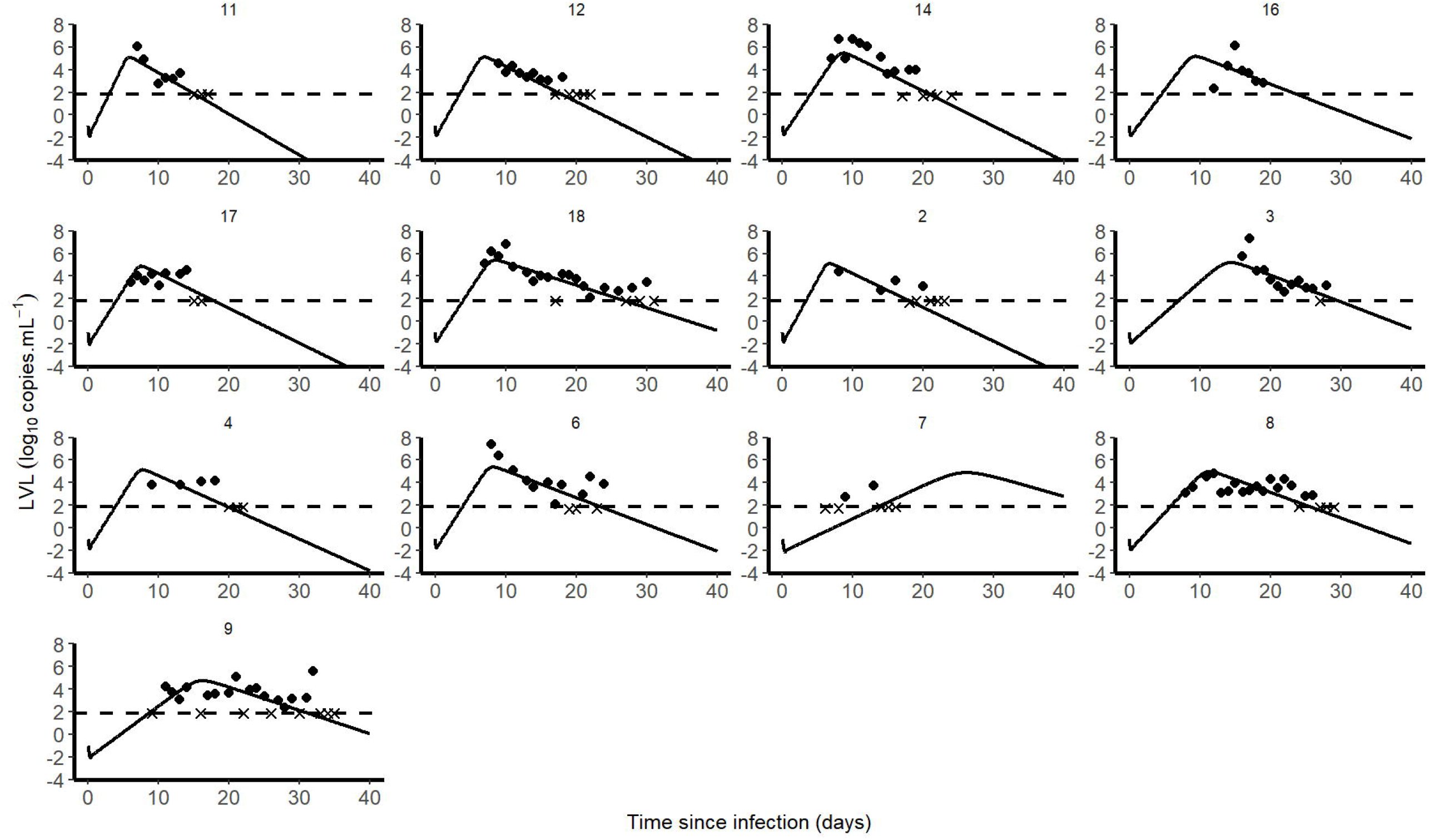
Individual predictions of SARS-CoV-2 of 13 untreated patients from Young et al. ^*3*^.

Our model along with the parameter estimates given above also inform us both on the time to initiate antiviral treatment, and the level of efficacy that needs to be achieved to reduce viral load ^6^. As limited information is available on the mechanisms leading to viral clearance, and how they may be modulated by treatment, we used our model to predict the effects of treatment at day 5 post symptoms, which corresponds to the time the viral load tends to peak in the absence of treatment in these data ^3^. We considered a simple case where the drug effectiveness is assumed to be constant after therapy initiation (see methods) and we calculated the minimal efficacy that would be needed to generate more than 2 logs of viral decline at peak viral load in the 13 studied patients (Fig. 2). As predicted by viral kinetic modeling theory ^2^, we found that the impact of treatment on peak viral load is inversely correlated with the time of treatment initiation. For a putative treatment blocking the viral production p and initiated at the time of infection, symptom onset, or 3 days post symptom onset, a median efficacy of at least 60, 90 and 99% in reducing viral replication would be needed, respectively, to generate more than 2 log of decline in the peak viral load (Fig. 2). The results obtained assuming 2 or 10 days of incubation are presented in Fig. S2 and S3. We also considered the case of a drug like hydroxychloroquine blocking viral infection (parameter β in Eq. 1). Results were similar to those obtained before as along as the treatment was initiated before or at symptom onset. However, initiating a treatment 3 days post-symptoms onset could not reduce peak viral load, regardless of the drug effectiveness (see Fig. S4).

**Figure 2:**
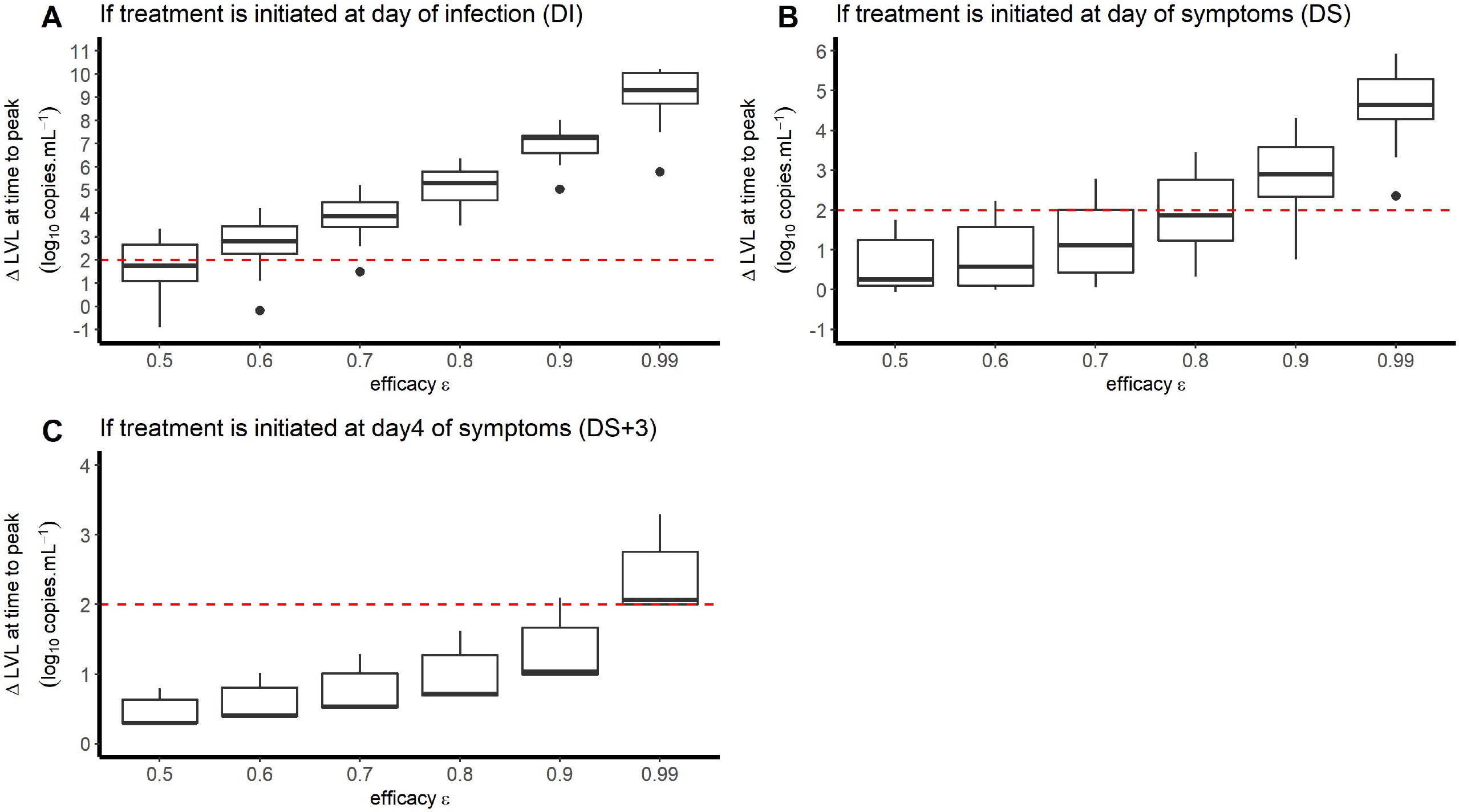
Reduction in viral load at day 5 post symptom onset according to the level of antiviral effectiveness of a treatment blocking the viral production and the timing of treatment initiation (A: at time of infection; B: at time of symptom onset; C: 3days after symptom onset). We assumed an incubation period of 5 days

The model including an innate immune response did not improve the data fitting and therefore was not selected for inclusion in the main text. However, the same analysis on viral dynamics and treatment was conducted and the conclusions remained unchanged (see Fig. S5, S6 and Table S2).

How do these levels of effectiveness compare with the antiviral drugs that are currently being investigated? To study this question, we assumed that the treatment antiviral effectiveness at time *t* after treatment initiation, ε(*t*), was related to the plasma total drug concentration 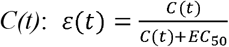 (except for remdesivir, see methods) and the mean antiviral effectiveness during the first 7 days of treatment is given by 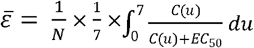. Given their pharmacokinetic and pharmacodynamic properties (Table 1), we calculated a mean antiviral effectiveness of up to 66% for lopinavir/ritonavir, 6% for hydroxychloroquine, 18% for IFN-β-1a and 87% for remdesivir. Given these estimates, these compounds are unlikely to have a dramatic effect on peak viral load if administered after the onset of symptoms. In fact, the effective concentrations will presumably be lower in patients, as drug availability may be further limited by protein binding (in particular for lopinavir, which has a protein binding rate > 98%) or capability to penetrate respiratory compartments. Importantly, levels of antiviral efficacy of ∼50% could nonetheless be relevant in a prophylactic setting, before symptom onset, to reduce viral replication in the upper respiratory tract and reduce the risk of large infiltration to the lung before an effective immune response is mounted to clear virus ^2^ Note, above we calculated the effectiveness of drugs administered in monotherapy for their usual dosing regimen. We did not consider drugs that could directly target infected cells and lead to their elimination, such as some monoclonal antibodies.

## Discussion

Overall our results emphasize that the PK/PD properties of lopinavir/ritonavir, hydroxychloroquine, IFN-β-1a and remdesivir make them unlikely to have a dramatic impact on nasopharyngeal viral load kinetics if they are administered after symptom onset. However, these drugs may be relevant to reduce viral replication if administered early, i.e., as a pre- or post-exposure prophylaxis. This conclusion however depends on a number of hypotheses that we discuss below.

First we focused on the capability of drugs to reduce the peak viral load making the implicit hypothesis that reducing peak viral load would likely reduce symptoms and disease severity. However the relationship between viral kinetics and disease severity is still debated, with several studies suggesting that viral load did not differ between patients with severe and non-severe disease ^21,22^. Related to this question, our study had some limitations. Our calculations relied on blood or plasma drug concentrations. Except for hydroxychloroquine, for which the ratio of lung to plasma concentration is known to be high^23,24^, the lung exposure of the other drugs that we considered is unknown, and their effect on viral load in the lower respiratory tract, as measured from broncho-alveolar aspirates for instance, may differ. In addition, the drug EC_50_ that we used were determined on Vero E6 cells, an *in vitro* system that may not reflect the *in vivo* EC_50_. For instance, we found in another study that hydroxychloroquine had no antiviral activity in a more physiological model of reconstituted human airway epithelium, and this may explain the absence of antiviral of activity of HCQ *in vivo* against SARS-CoV-2^24,25^. Finally we focused solely on the antiviral effects of these drugs, and did not consider other potential effects of these drugs, such as their immunomodulatory effects. Such effects have been suggested for drugs that are not purely antivirals, such as hydroxychloroquine and IFN-β-1a^26,27^.

Another implicit implication of our work is the benefit of drugs used for prophylaxis, i.e., before exposure to the virus. In that case the objective of the treatment may be to “flatten the peak viral load” (by analogy with the now popular terminology of epidemiological models) but also to prevent infection. Our deterministic, ODE-based model, cannot reproduce virus extinction, but this can be captured using stochastic version of the same model^28^.

Our modeling provides estimates of viral kinetic parameters, in particular R_0_ and the loss rate of infected cells, *δ*, and the limit of extrapolation of these parameters also need to be well understood. The advantage of studying this series of patients was the fact that viral load was sampled extremely frequently and early in the infection. However the number of patients we studied was quite small (only 13), and the patients were rather young (37 years, range 31-56 years) and may not be representative of patients that are typically evolve to have severe forms of the disease ^21^. Therefore, more data will be needed, from various populations, to estimate precisely the parameter distribution in the population of patients that are most in need of antiviral therapy. Finally, our estimate of the loss rate of free virus and infected cells are constant over time, which neglects the effects of the adaptive immune response. For instance antibodies, that emerge in the second week after symptom onset^29,30^, may contribute to accelerate viral clearance or reduce viral infectivity. However, to include these effects would require more complex models and quantitative data on these antibodies and their in vivo effects, which are currently lacking. Further, our analysis of the antiviral effects of repurposed drugs focused on their ability to reduce the peak viral load, which typically occurs well before the antibody response emerges and hence the analysis we presented here should not be affected in any major way by our neglect of antibody responses.

Future models of drug efficacy may need to account for viral resistance, as it is possible that continued viral replication in the presence of drug will select for drug resistant mutations ^31^, although coronaviruses are unusual in that they appear to have low mutation rates due to RNA proofreading capability^32^. Drug combination therapy and more aggressive dosing, including consideration of loading doses to rapidly achieve therapeutic exposures, may be beneficial to maximize efficacy of these repurposed antiviral agents^33^. For all these reasons, the outcome of randomized clinical trials remains urgently needed, and the analysis of their impacts not only on viral clearance but also on disease severity will be critical to design more potent drugs.

## Data Availability

Data were originally provided in Young et al. doi:10.1001/jama.2020.3204 "where written informed consent was obtained from study participants for collection of biological samples after review and approval of the study protocol by the institutional ethics committee"

https://jamanetwork.com/journals/jama/fullarticle/2762688

## Study highlights (total answers should be less than 150 words)

- What is the current knowledge on the topic?

Repurposed drugs are being evaluated in clinical trials but little is known about their efficacy on SARS-CoV-2 viral kinetics.

- What question did this study address?

Our study aims to combine PK/PD and viral kinetics modelling to anticipate the effects of lopinavir/ritonavir, hydroxychloroquine, IFN-β-1a and remdesivir,

- What does this study add to our knowledge?

Given the predicted efficacy of lopinavir/ritonavir, hydroxychloroquine, IFN-β-1a and remdesivir, it is unlikely that these drugs will have a major effect on viral kinetics if they are administered as monotherapy after symptom onset.

- How might this change drug discovery, development, and/or therapeutics?

Our results suggest that these drugs should be evaluated in persons exposed to the virus but prior to appearance of the first symptoms.

## Author contributions

AG, RK, ASP and JG designed the research, analyzed viral kinetics data and wrote the manuscript. JB, EC and PS performed PK/PD predictions; AP, OT and MRC performed in vitro experiments. XDL, FM and DM provided guidance on clinical and virological implications.

## Notes

**Conflict of interest** Jérémie Guedj has consulted for F. Hoffman-La Roche. All other authors declared no competing interests for this work.

### Competing Interest Statement

The authors have declared no competing interest.

### Funding Statement

Antonio Gonçalves was funded by a grant from Roche Pharmaceutical Research and Early Development.
Portions of this work were done under the auspices of the U.S. Department of Energy under contract 89233218CNA000001. We also gratefully acknowledge the support of the U.S. Department of Energy through the LANL/LDRD Program for this work as well as N.I.H. grants R01 AI028433, R01 OD011095, R01 AI078881 and P01 AI131365 (ASP).

## References

1. Cao, B. et al. A Trial of Lopinavir–Ritonavir in Adults Hospitalized with Severe Covid-19. N. Engl. J. Med. 382, 1787–1799 (2020).

2. Friberg, L. E. & Guedj, J. Acute bacterial or viral infection—What’s the difference? A perspective from PKPD modellers. Clin. Microbiol. Infect. (2019). doi:10.1016/j.cmi.2019.12.008

3. Young, B. E. et al. Epidemiologic Features and Clinical Course of Patients Infected With SARS-CoV-2 in Singapore. JAMA (2020). doi:10.1001/jama.2020.3204

4. Zou, L. et al. SARS-CoV-2 Viral Load in Upper Respiratory Specimens of Infected Patients. N. Engl. J. Med. 382, 1177–1179 (2020).

5. Lauer, S. A. et al. The Incubation Period of Coronavirus Disease 2019 (COVID-19) From Publicly Reported Confirmed Cases: Estimation and Application. Ann. Intern. Med. (2020). doi:10.7326/M20-0504

6. Best, K. et al. Zika plasma viral dynamics in nonhuman primates provides insights into early infection and antiviral strategies. Proc. Natl. Acad. Sci. 114, 8847–8852 (2017).

7. Banerjee, S., Guedj, J., Ribeiro, R. M., Moses, M. & Perelson, A. S. Estimating biologically relevant parameters under uncertainty for experimental within-host murine West Nile virus infection. J. R. Soc. Interface 13, (2016).

8. Baccam, P., Beauchemin, C., Macken, C. A., Hayden, F. G. & Perelson, A. S. Kinetics of influenza A virus infection in humans. J. Virol. 80, 7590–7599 (2006).

9. Muus, C. et al. Integrated analyses of single-cell atlases reveal age, gender, and smoking status associations with cell type-specific expression of mediators of SARS-CoV-2 viral entry and highlights inflammatory programs in putative target cells. bioRxiv (2020). doi:10.1101/2020.04.19.049254

10. Agostini, M. L. et al. Coronavirus Susceptibility to the Antiviral Remdesivir (GS-5734) Is Mediated by the Viral Polymerase and the Proofreading Exoribonuclease. mBio 9, (2018).

11. Gonçalves, A., Mentré, F., Lemenuel-Diot, A. & Guedj, J. Model Averaging in Viral Dynamic Models. AAPS J. 22, 22–48 (2020).

12. Pawelek, K. A. et al. Modeling within-host dynamics of influenza virus infection including immune responses. PLoS Comput. Biol. 8, e1002588 (2012).

13. Wang, K. et al. Integrated Population Pharmacokinetic/Viral Dynamic Modelling of Lopinavir/Ritonavir in HIV-1 Treatment-Naïve Patients. Clin. Pharmacokinet. 53, 361–371 (2014).

14. Morita, S., Takahashi, T., Yoshida, Y. & Yokota, N. Population Pharmacokinetics of Hydroxychloroquine in Japanese Patients With Cutaneous or Systemic Lupus Erythematosus: Ther. Drug Monit. 38, 259–267 (2016).

15. Hu, X. et al. COMPARE: Pharmacokinetic profiles of subcutaneous peginterferon beta-1a and subcutaneous interferon beta-1a over 2 weeks in healthy subjects: Pharmacokinetics of peginterferon beta-1a and s.c. interferon beta-1a. Br. J. Clin. Pharmacol. 82, 380–388 (2016).

16. Francisco, E. M. EMA provides recommendations on compassionate use of remdesivir for COVID-19. Eur. Med. Agency (2020). <https://www.ema.europa.eu/en/news/ema-provides-recommendations-compassionate-use-remdesivir-covid-19> Accessed 10 May 2020.

17. Pizzorno, A. et al. Characterization and treatment of SARS-CoV-2 in nasal and bronchial human airway epithelia. (2020). doi:10.1101/2020.03.31.017889

18. Smith, A. M. Host-pathogen kinetics during influenza infection and coinfection: insights from predictive modeling. Immunol. Rev. 285, 97–112 (2018).

19. Miao, H., Xia, X., Perelson, A. S. & Wu, H. On identifiability of nonlinear ode models and applications in viral dynamics. SIAM Rev. Soc. Ind. Appl. Math. 53, 3–39 (2011).

20. Stafford, M. A. et al. Modeling plasma virus concentration during primary HIV infection. J. Theor. Biol. 203, 285–301 (2000).

21. Zheng, S. et al. Viral load dynamics and disease severity in patients infected with SARS-CoV-2 in Zhejiang province, China, January-March 2020: retrospective cohort study. BMJ 369, (2020).

22. Liu, S., Zhou, B., Valdes, J. D., Sun, J. & Guo, H. Serum Hepatitis B Virus RNA: A New Potential Biomarker for Chronic Hepatitis B Virus Infection. Hepatology 69, 1816–1827 (2019).

23. Yao, X. et al. In Vitro Antiviral Activity and Projection of Optimized Dosing Design of Hydroxychloroquine for the Treatment of Severe Acute Respiratory Syndrome Coronavirus 2 (SARS-CoV-2). Clin. Infect. Dis. (2020).doi:10.1093/cid/ciaa237

24. Maisonnasse, P. et al. Hydroxychloroquine in the treatment and prophylaxis of SARS-CoV-2 infection in non-human primates. Research Square (2020).doi:10.21203/rs.3.rs-27223/v1

25. Boulware, D. R. et al. A Randomized Trial of Hydroxychloroquine as Postexposure Prophylaxis for Covid-19. N. Engl. J. Med. 0, null (2020).

26. Liu, J. et al. Hydroxychloroquine, a less toxic derivative of chloroquine, is effective in inhibiting SARS-CoV-2 infection in vitro. Cell Discov. 6, 1–4 (2020).

27. Ingraham, N. E. et al. Immunomodulation in COVID-19. Lancet Respir. Med. (2020).doi:10.1016/S2213-2600(20)30226-5

28. Czuppon, P. et al. Predicted success of prophylactic antiviral therapy to block or delay SARS-CoV-2 infection depends on the targeted mechanism. (2020).doi:10.1101/2020.05.07.20092965

29. Woelfel, R. et al. Clinical presentation and virological assessment of hospitalized cases of coronavirus disease 2019 in a travel-associated transmission cluster. medRxiv (2020).doi:10.1101/2020.03.05.20030502

30. Wu, F. et al. Neutralizing antibody responses to SARS-CoV-2 in a COVID-19 recovered patient cohort and their implications. medRxiv (2020).doi:10.1101/2020.03.30.20047365

31. Perelson, A. S., Rong, L. & Hayden, F. G. Combination Antiviral Therapy for Influenza: Predictions From Modeling of Human Infections. J. Infect. Dis. 205, 1642–1645 (2012).

32. Denison, M. R., Graham, R. L., Donaldson, E. F., Eckerle, L. D. & Baric, R. S. Coronaviruses. RNA Biol. 8, 270–279 (2011).

33. Smith, P. F., Dodds, M., Bentley, D., Yeo, K. & Rayner, C. Dosing will be a key success factor in repurposing antivirals for COVID[19. Br. J. Clin. Pharmacol. (2020).doi:10.1111/bcp.14314

